# Short-term Projections based on Early Omicron Variant Dynamics in England

**DOI:** 10.1101/2021.12.30.21268307

**Authors:** Matt J. Keeling, Ellen Brooks-Pollock, Rob Challen, Leon Danon, Louise Dyson, Julia R. Gog, Laura Guzmán Rincón, Edward M. Hill, Lorenzo Pellis, Jonathan M. Read, Michael J. Tildesley

## Abstract

Throughout the ongoing COVID-19 pandemic, the worldwide transmission and replication of SARS-COV-2, the causative agent of COVID-19 disease, has resulted in the opportunity for multiple mutations to occur that may alter the virus transmission characteristics, the effectiveness of vaccines and the severity of disease upon infection. The Omicron variant (B.1.1.529) was first reported to the WHO by South Africa on 24 November 2021 and was declared a variant of concern by the WHO on 26 November 2021. The variant was first detected in the UK on 27 November 2021 and has since been reported in a number of countries globally where it is frequently associated with rapid increase in cases. Here we present analyses of UK data showing the earliest signatures of the Omicron variant and mathematical modelling that uses the UK data to simulate the potential impact of this variant in the UK. In order to account for the uncertainty in transmission advantage, vaccine escape and severity at the time of writing, we carry out a sensitivity analysis to assess the impact of these variant characteristics on future risk.

## Introduction

Throughout the ongoing COVID-19 pandemic, the worldwide transmission and replication of SARS-COV-2, the causative agent of COVID-19 disease, has resulted in the opportunity for multiple mutations to occur that may alter the virus’ transmission characteristics, the effectiveness of vaccines and the severity of disease upon infection. Some of the variants that have emerged have been designated variants of concern (VOC) by the World Health Organisation (WHO) owing to the risks that they pose and the potential for impacting the effectiveness of the ongoing worldwide vaccination programme.

The Omicron variant (B.1.1.529) was first reported to the WHO by South Africa on 24 November 2021 and was declared a variant of concern by the WHO on 26 November 2021 [1]. The variant was first detected in the UK on 27 November 2021 [2] and has since been reported in a number of countries globally where it is frequently associated with rapid increase in cases [3]. The variant has around 60 mutations from the original wildtype variant from Wuhan with a large number of these (more than 30) in the spike protein [4]; these spike mutations have raised significant concerns regarding the potential effectiveness of existing vaccines and the degree of protection from past infections. The rapid spread of the Omicron variant within Gauteng province in South Africa, where it was first detected on 23 November 2021 [5], led to initial speculation that it had a significant transmission advantage over other circulating variants. At the time of first detection, less than 25% of adults in South Africa were fully vaccinated [6-7] and there was substantial uncertainty regarding how rapidly this variant would spread in other countries with higher vaccine uptake.

Prior to the introduction of the Omicron variant into the UK, the number of daily reported cases in the UK was high but relatively constant whilst the average number of daily hospital admissions and deaths as a result of COVID-19 infections was falling throughout November 2021. However, overall hospital occupancy has remained high, with the National Health Service running close to capacity. It therefore became crucial to develop a rapid but comprehensive understanding of characteristics of this VOC and the impact of large-scale spread of the variant upon hospital admissions and deaths in subsequent weeks. Additionally, whilst around 80% of individuals aged 12 or over in the UK have received two doses of vaccine, the early data indicating that the Omicron variant was exhibiting some level of vaccine escape, combined with the presence of waning immunity in the months following the second dose, suggested that there was an urgent need to accelerate the booster vaccination campaign in order to increase protection levels across the population.

Three aspects of the Omicron variant are potentially different to Delta: its transmissibility; the degree of immune escape; and its severity. The higher growth of Omicron than Delta in South Africa suggests that a combination of higher transmissibility and immune escape (from vaccination and/or prior infection) results in a higher growth rate than Delta [8]. Antibody neutralisation studies indicate lower neutralisation compared to Delta of both convalescent and vaccinated sera, suggesting immune escape [9-11]. These studies are also supported by South African data showing high reinfection rates compared to Delta [12], and early UK data of PCR positive individuals, which indicate lower vaccination efficacies of both AstraZeneca and Pfizer vaccinations [4]. It has been claimed that Omicron displays decreased severity compared to Delta, as the South African case-hospitalisation ratio appears to be lower for the emerging Omicron wave than for Delta. However, these data are triply confounded by (i) higher vaccination rates and prior infections at the time of the Omicron wave compared to the Delta wave; (ii) time lags before hospitalisations and deaths; and (iii) different age distributions of infections between the two waves. Analysis of very early UK data did not find a significant difference between Omicron and Delta hospitalisation rates when controlling for other factors, although with only 24 Omicron hospitalisations in the dataset studied, it is early to draw firm conclusions from this [13]. There are no credible claims of higher severity for Omicron compared to Delta, and therefore in this work we consider a range of relative severities from 10% to 100%.

In this paper, we present analyses of UK data showing the earliest signatures of the Omicron variant and mathematical modelling that uses the UK data to simulate the potential impact of this variant in the UK. We utilise a previously developed mathematical model [14-15] that simulates the spread of SARS-CoV-2 within the seven NHS regions of England to predict the future spread of the Omicron variant, as well as the number of daily hospital admissions and deaths as a result of infection and how that spread may depend upon the introduction of non-pharmaceutical interventions. In order to account for the uncertainty in transmission advantage, vaccine escape and severity at the time of writing, we carry out a sensitivity analysis to assess the impact of these variant characteristics on future risk.

### Early signatures of Omicron transmission in UK Data

Because of the high numbers of cases in the UK during November 2021, early emergence of cases infected with the Omicron variant were not immediately evident in the aggregate case data. We therefore explored the UKHSA ‘linelist’ case data for early signs of Omicron variants transmission. Omicron variants are further subdivided into sublineages BA.1, BA.2 and BA.3, with BA.1 by far the most prevalent variant of the three. Fortunately, BA.1 and BA.3 exhibit a deletion in the spike protein at position S69:70, a deletion also exhibited by the Alpha variant, which causes it to fail to amplify in the TaqPath S-Gene target (S-Gene Target Failure, SGTF) employed in UK routine testing ‘Lighthouse’ laboratories. BA.2 does not display SGTF, and so cannot be discerned in this way, however so far very few sequenced cases of BA.2 have been found in the UK.

It is therefore possible to track the progression of Omicron in routine tests that show SGTF, assuming that the majority of these are caused by Omicron cases. Figure 1 shows the coverage of laboratories able to identify SGTF. Coverage by Lighthouse laboratories varies in space and time. The highest coverage (over 80% of all tests) was in North East England in early December 2021, the lowest coverage was in Southwest England with less than a quarter of samples tested for SGTF, while the coverage for London is spatially patchy. This suggests that early indicators from the North East are likely to be more accurate than other regions and a possible underestimate of Omicron case burden, especially in the South West of England (Figure 1).

**Figure 1:**
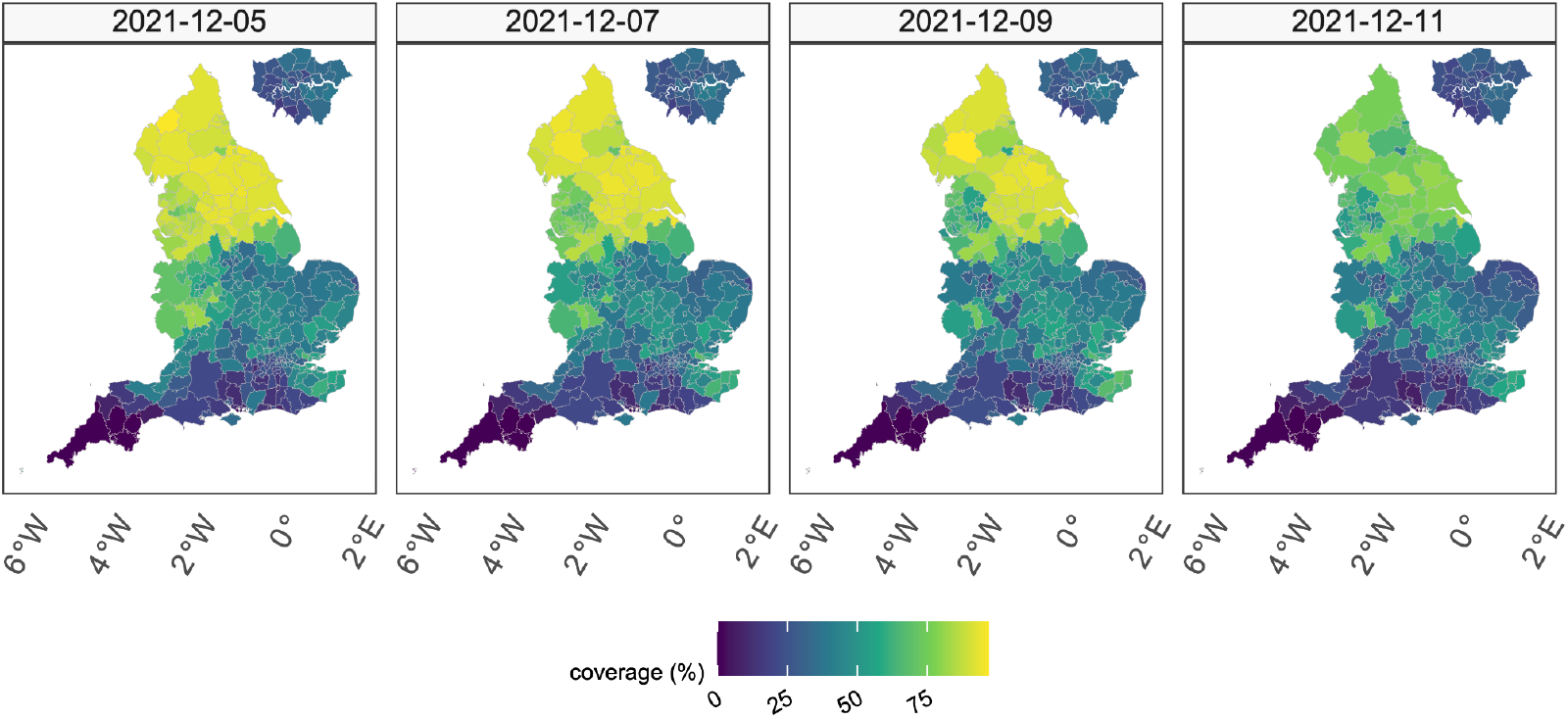
Coverage of testing in England able to identify S-Gene Target Failure (SGFT). Regions of the South West which have lower coverage, are less likely to be able to rapidly discriminate between Omicron and Delta variants.

Aggregating testing data across all age groups and all regions in England, we observed a rapid increase in the proportion of cases that display SGTF (compared to all of those where presence or absence of the S-gene target was determined), starting towards the end of November 2021 (Figure 2A). By considering the proportion of cases with SGTF (Figure 2A), rather than the number of cases with SGTF, we are able to consider the most recent data as reporting delays have a minimal effect on the proportion (assuming SGTF and non-SGTF have the same distribution of delays). Figure 2B illustrates the absolute incidence of S-gene positive and SGTF cases during the period 1 November to 13 December 2021: without S-gene detection the number of SGTF cases would have been too small to identify any growth at this early stage. During this period, the S-gene positive cases (largely the Delta variant) showed a slow decline, consistent with high levels of population immunity, whereas the SGTF cases showed a rapid increase (Figure 2C). Although still very large and positive, the growth rate of SGTF cases (and S-gene positive cases) may have declined slightly since the start of December, although this could be attributed to delays in reporting or changes in the age-distribution of cases. (We note that the proportion of cases that display SGTF must eventually saturate at close to 100%, so the growth rate of this proportion only provides useful information when Omicron is relatively uncommon.)

**Figure 2.**
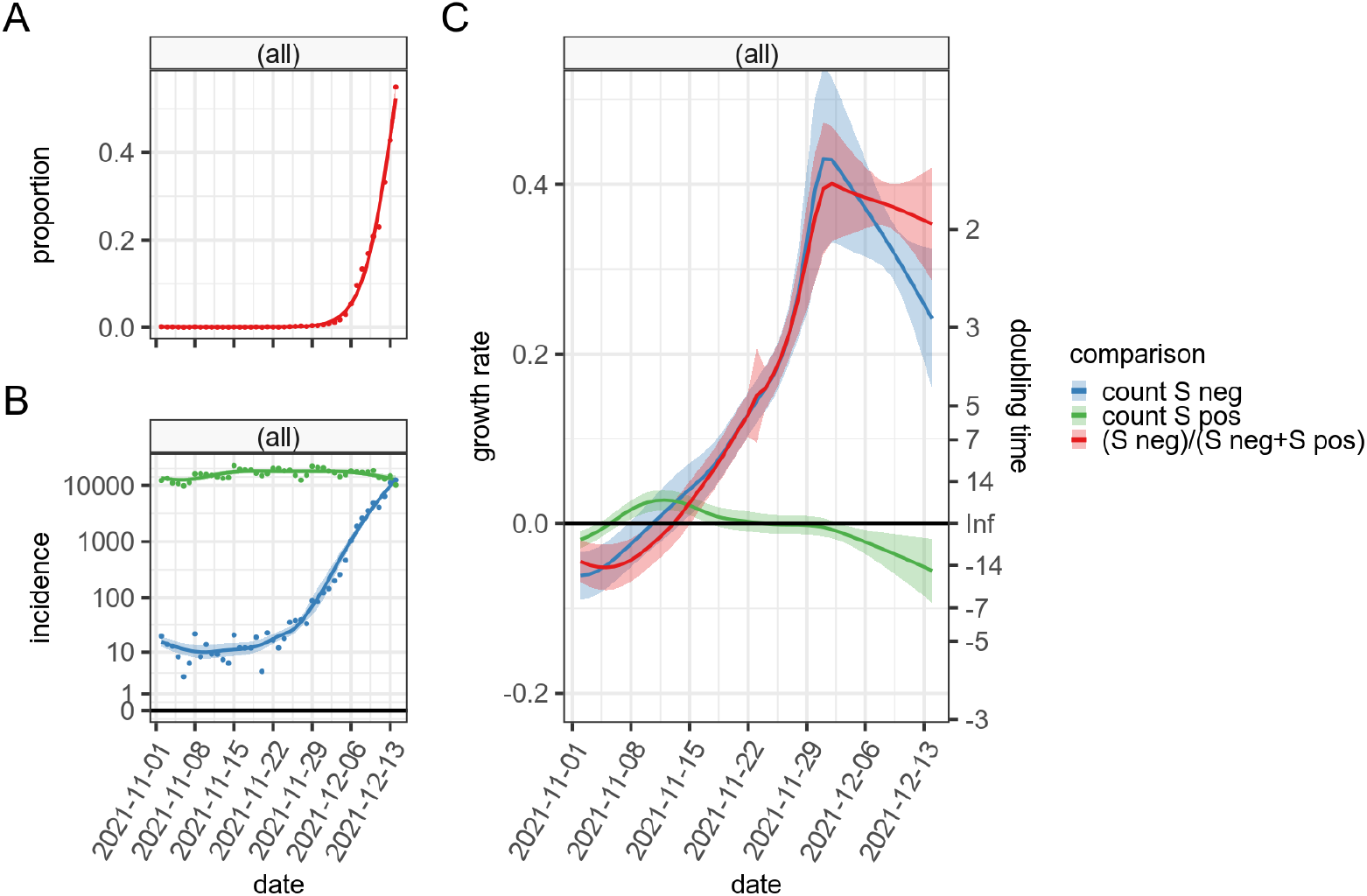
S-gene target failure (SGTF) in the data. A) The proportion of all test positive cases in the UK with SGTF between 1 November 2021 and 13 December 2021. B) The absolute incidence (new cases identified per day) of cases with S-gene target failure (blue) and S-gene detection (green). C) The inferred growth rates of S-gene positive (green) and SGTF cases (blue) and the ratio between them (red).

### COVID cases that are confirmed reinfections

Since higher reinfection rates have been observed for Omicron than Delta [12], another indicator of the scale and speed of the omicron wave, as well as the extent of vaccine escape, is the proportion of cases that are reinfections. the signal of changing patterns in reinfections give a separate view of the unfolding Omicron wave, also partial and imperfect; reinfection information alone cannot discriminate between delta and omicron variants, but can be useful as an additional ‘indicative’ measure of omicron infection dynamics, and a signal of the likely degree of cross-protection from previous infections.

The UKHSA ‘linelist’ case data is limited to individuals in England who tested positive for SARS-CoV-2 for the first time only which, if there is substantial reinfection, underestimates incidence. Reinfection data provided by UKHSA is limited to infections which occur more than 90 days following the last positive test. We modelled the number of reinfections in both Pillar 1 and 2 using a generalized additive model with a penalised spline for secular time and accounting for the total number of samples tested (first + reinfections) with an offset term. Reinfections were occurring at a relatively low rate (1-3%) across all age groups throughout September-November 2021, and are likely to be reinfection with Delta (Figure 3). During early December 2021, we see a significant increase in the reinfection rate in nearly all age groups – but a particularly rapid increase in young adults (20-29 year olds) – suggesting a change in population immunity to the dominant variants, and likely indicative of omicron invasion and transmission establishment.

**Figure 3.**
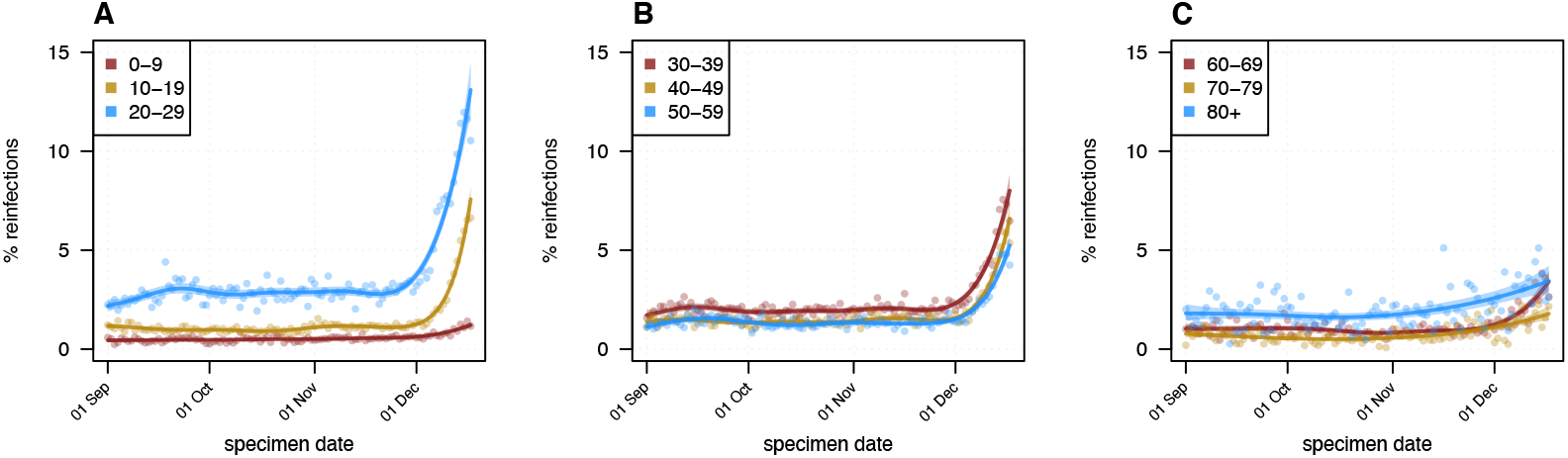
Estimated daily % of cases that are reinfections for England, stratified by age group, showing substantial increases during December in many age groups above a previously relatively constant reinfection rate for each age group, indicative of the omicron invasion. Observations are shown as dots, fitted spline as lines and 95% confidence intervals as shaded regions.

Stratifying by age group and region of England (Figure S2) highlights the rapid rise in reinfection in the 20-29 age group is seen in all regions including regions with low SGTF coverage, and is not just confined to London. Reinfection rates are rising in most age groups in most regions, particularly adults aged 20 to 60.

### Epidemiological characteristics of the Omicron Variant

One of the major challenges with projecting the potential dynamics of Omicron is the limited amount of data to inform critical transmission and disease parameters. Because of the recent emergence of the Omicron variant there is still little data on vaccine efficacy against severe disease, including hospital admissions or death. The impact of vaccination on onward transmission if an individual becomes infected has been poorly quantified for any variant throughout the pandemic due to difficulty in accurately measuring secondary attack rates (although some information is available from detailed household studies). There is also limited information on the severity of Omicron compared to Delta in terms of the relative risk of hospital admission, and the duration of hospital stays. Each of these factors can have a profound effect on the projected scale of the outbreak, the impact on health services, and the impact of control measures.

As described above, although genotyping following PCR detection is the ideal gold-standard for confirming Omicron infection, the rapid increase in Omicron cases since its arrival in the UK in late November 2021 can be readily observed as an increase in test positive cases with S-gene target failure (SGTF). This approach has previously been successfully used to monitor the early growth of the Alpha and Delta variants [16-18]. Figure 4 shows the growth in the proportion of PCR tests that are S-gene negative (relative to all those that generate a clear signal from the TaqPath testing), compared to the proportion of infections that are Omicron from simulations.

**Figure 4:**
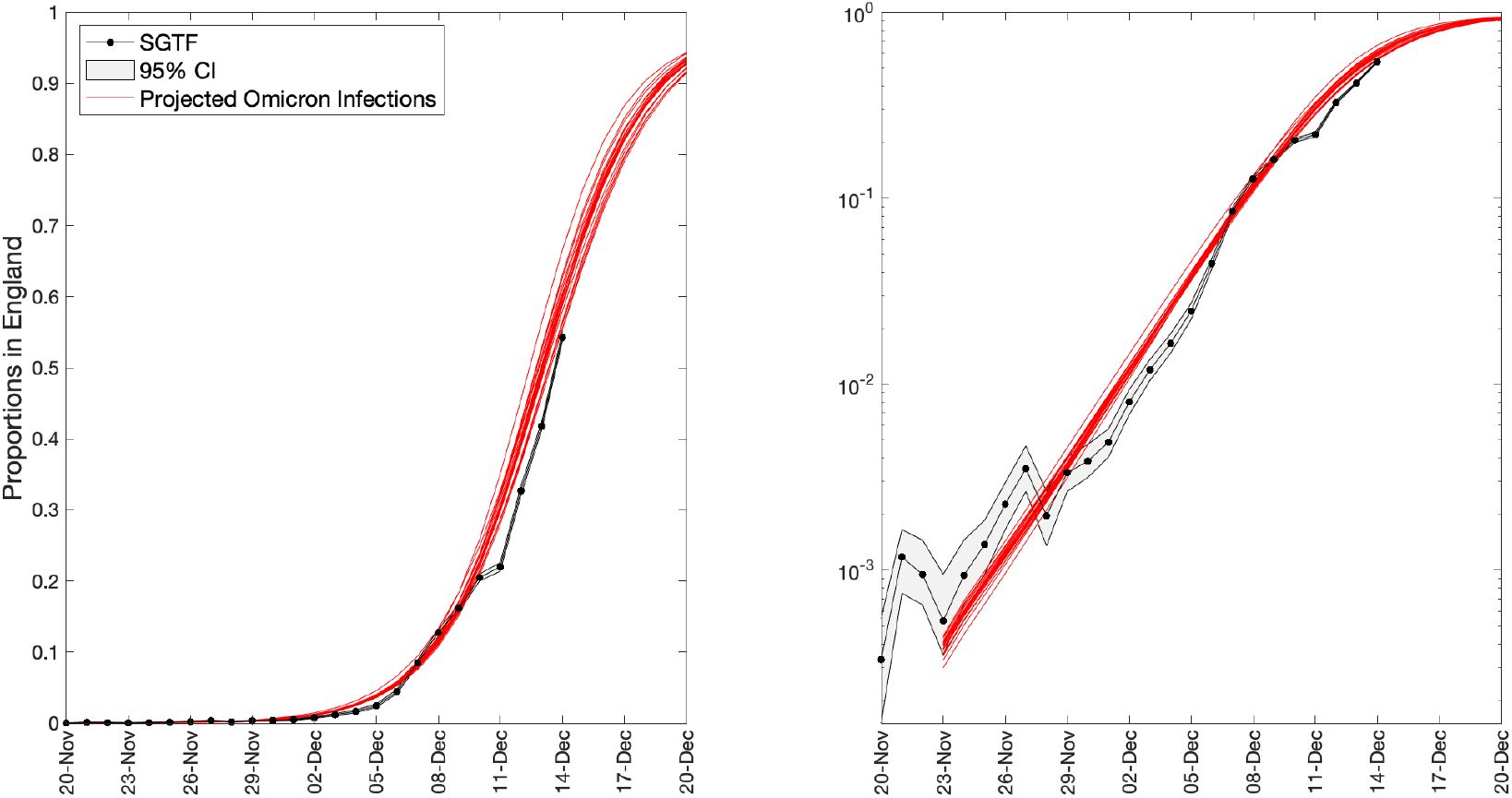
Comparison of results from the S-gene target failure within the TaqPath PCR detection process (black) with simulation results (red). The S-gene data show the proportion of tests with a strong PCR signal (cycle threshold less than 30) that are negative for the S-gene target (black dots show the proportion, while grey bars are the associated 95% confidence intervals). The simulation results are the projected proportion of newly infected individuals that have the Omicron variant. The left-hand figure is on a linear scale, while the right-hand figure is on a logarithmic scale.

Figure 4 is a visual comparison between the rise of the SGTF cases in the data and the rise of Omicron infections from our transmission model. When matching model results and data (as we do for parameter inference), we must include the distribution of delays between infection and testing, the age-dependent biases in the likelihood of performing a test, and the low level of S-gene negative tests that were present before the invasion of Omicron. However, here we simply plot here the most parsimonious results from the model projections: the projected proportion of new infections that are due to the Omicron variant.

Achieving the fit to SGTF growth (Figure 4) requires a set of assumptions about the vaccine efficacy against Omicron and the level of cross-protection against Omicron from previous infections (see below). Once these are set, we utilise our existing MCMC inference machinery to determine the relative level of Omicron transmission compared to Delta. To achieve the high growth rate of S-gene negative results observed in all regions, we infer Omicron to be 3.18 (95% credible interval: 2.95 - 3.78) times more transmissible than Delta, assuming the same generation time for both variants.

Although we have seen an increase in reinfection, particularly in those aged 20-38 and in London (Figure 3 and Figure S2), it is difficult to use this information to directly estimate the level of cross protection afforded by infection with previous variants. The pattern of reinfection is particularly difficult to assess due to extreme heterogeneities in testing patterns: those regularly taking lateral flow tests are most likely to detect (potentially mild or asymptomatic) reinfection. In our modelling results we assume that those previously infected (whose immunity has not waned) have 90% immunity to infection by Omicron (compared to 100% against other variants), while protection against hospital admission is higher again at 95% - such that previous infections provide an additional level of protection against severe disease.

Capturing vaccine efficacy is more complex, as this depends on the type of vaccine received, the number of doses, the age of the individual and the time since vaccination (and potentially the time between doses). Here we compare early estimates of vaccine efficacy for Omicron from UKHSA [4] with simple extrapolations from the model structure (Figure 5). In our model framework (see Supplementary methods), we assume those recently vaccinated have a fixed vaccine efficacy across all age-groups that depends only on the type of vaccine given, with mRNA vaccines (Pfizer and Moderna) assumed to generate equal efficacy. Waning of vaccine efficacy is modelled as a two-step process with protection (against infection) eventually dropping to zero. Booster vaccination moves all vaccinated individuals (even those that have waned) to a boosted class, with generally higher efficacy. The initial level of protection against the Delta variant, the protection offered by the booster and the speed of waning are matched to estimated values (coloured and black squares in Figure 5). Recent data on projection against the Omicron variant (grey circles in Figure 5), hints at a potentially simple relationship:

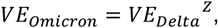

where, Z=4 provides a good agreement for the efficacy against symptomatic infection (comparing blue and grey circles) - this generates an efficacy against symptomatic infection with Omicron of 71% after boosting. We extend this scaling to estimate the protection against hospital admissions for Omicron (shown in red); this generates an efficacy against severe disease with Omicron of 85% after boosting.

**Figure 5:**
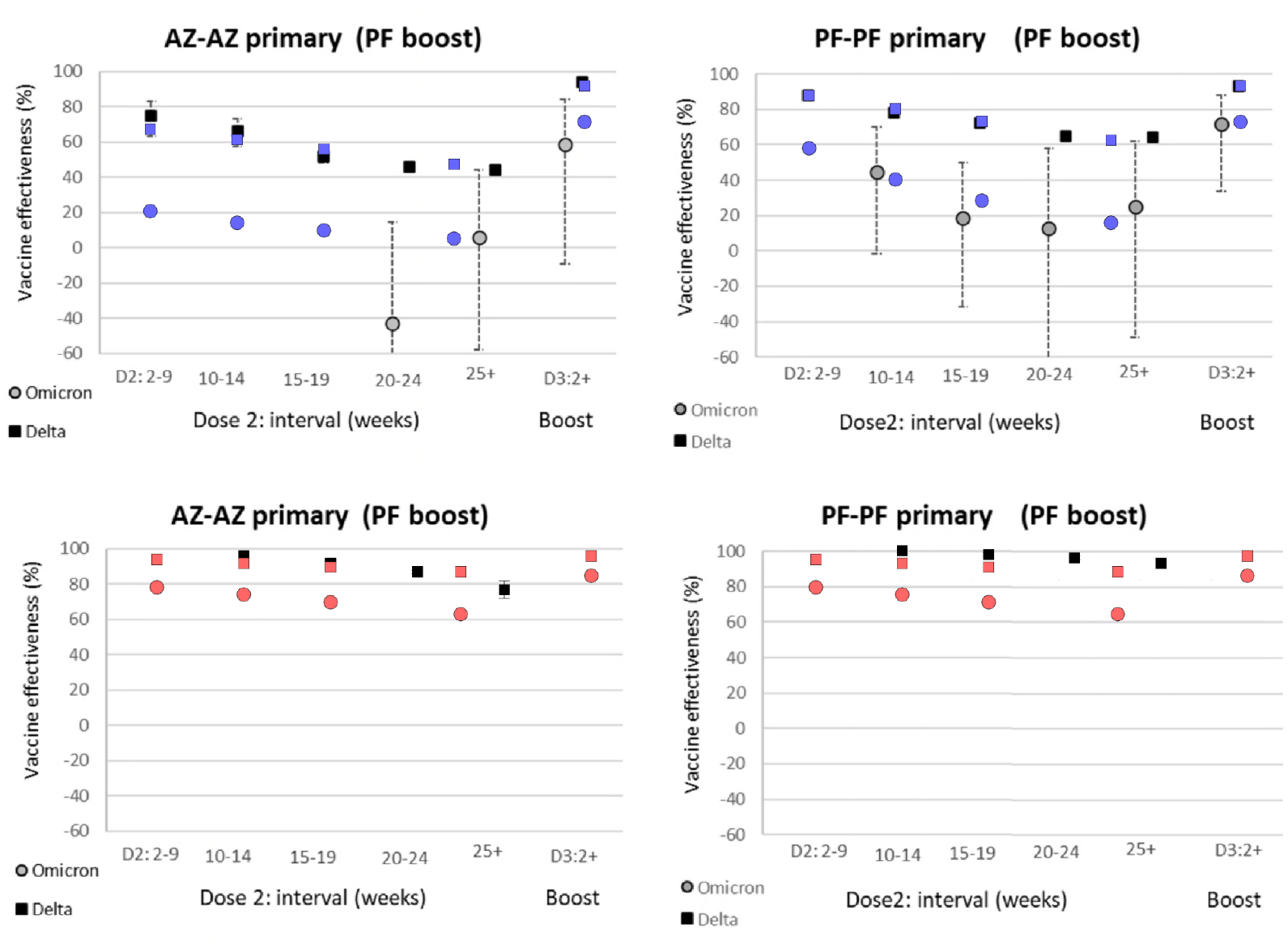
Vaccine efficacy against symptomatic infection (top panels) and hospital admissions (lower panels), showing results estimated by UKHSA (black) and values used in the projections (colours). Efficacies for Delta and Omicron are shown as squares and circles respectively.

The assumptions about vaccine efficacy and protection from previous infection with other variants have direct implications about which age-groups suffer the greatest burden of the Omicron wave based on the current level of immunity in the population (Figure 6). Older individuals are protected by vaccine efficacy from boosters (although the rollout of boosters is now reaching other age-groups); whereas younger individuals are more likely to be protected by past infection (or a mixture of past infection combined with vaccination). Therefore, if vaccine efficacy is lower than estimated then infection will be more focused in the older age-groups; if protection from past infection is lower, then infection will be primarily in the younger individuals.

**Figure 6:**
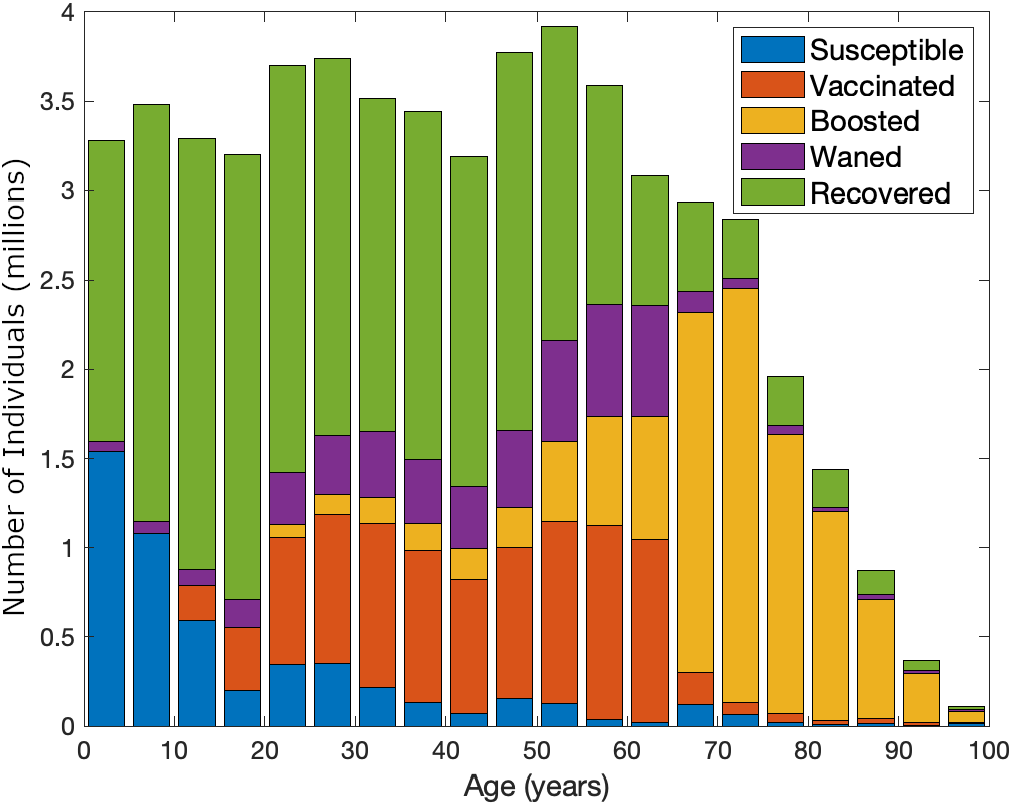
Estimated distribution of immune status by age from the model at the start of December 2021. Susceptible individuals (who have no protection, blue) are concentrated in the youngest age-groups, while those whose immunity is from infection (recovered in green) are generally below 60. For those above 60 years old, the vast majority are protected by booster vaccines (yellow).

A comparison of the age distribution of Delta and Omicron cases as determined by SGFT status (Figure 7 left-hand age-pyramid) reveals some interesting characteristics (echoing the results of Figure S1). Delta cases dominate in the younger age groups (5-9 and 10-14) and peak again in those 35-44 whilst Omicron cases are highest in those aged 20-29. In contrast, modelled infections (right-hand pyramid of Figure 7) show less heterogeneity, with Omicron infection equally distributed between ages 25 and 44. Part of this discrepancy may lie in the difference between infection and cases; infections only become reported cases if an individual takes a lateral flow or PCR test, which in turn depends upon disease symptoms or testing practices.

**Figure 7:**
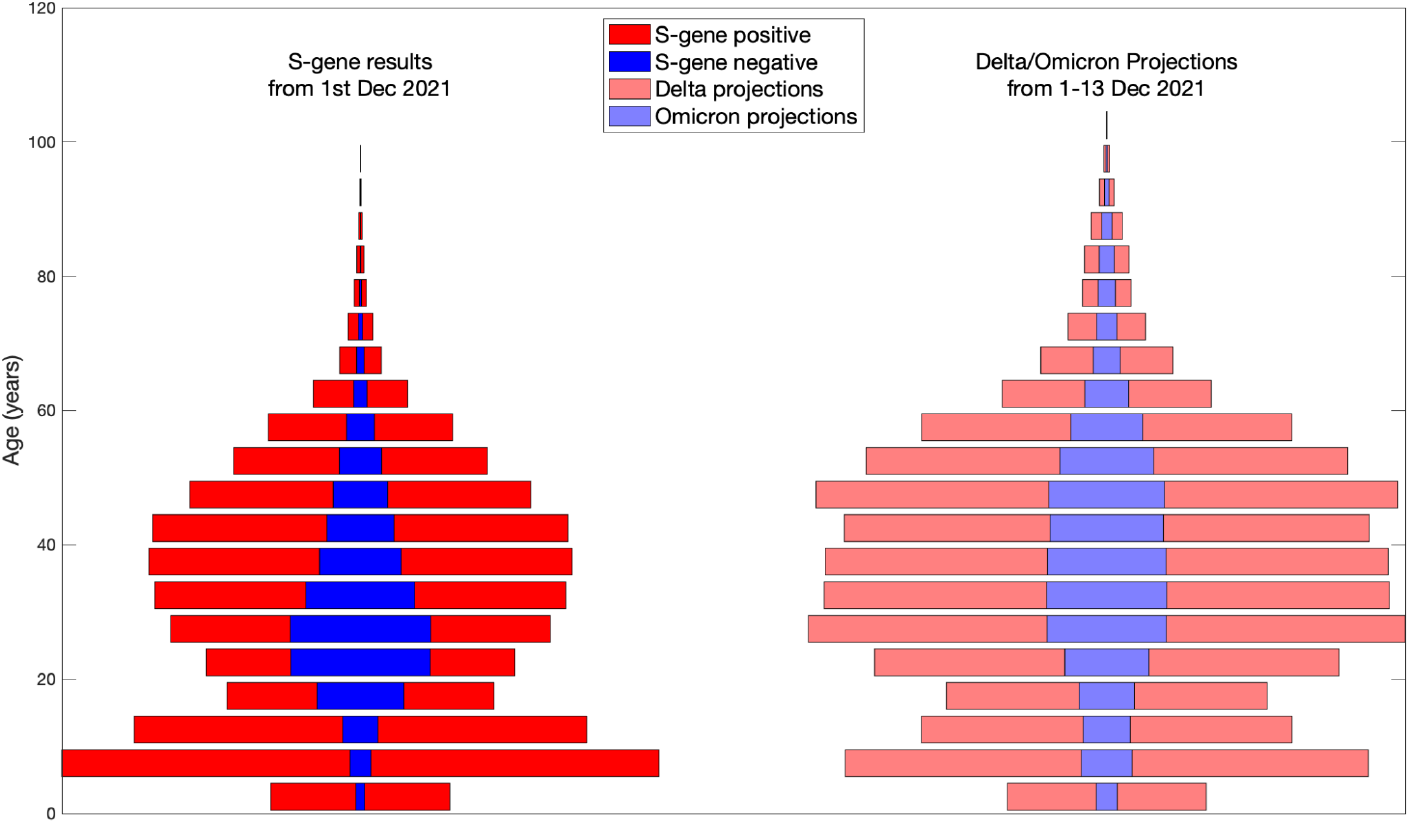
Comparison of age-structured data (left) and model projections (right) for S-gene target failure and variant type respectively. The age-bars are stacked such that in the left-hand plot, the total width of a bar is the number of all reliable samples with an S-gene target result (positive or negative), while red and blue correspond to samples that are S-gene positive (largely Delta) or S-gene negative (largely Omicron) respectively. For the model simulations, the number of infections from each variant can be computed; we note that the right-hand plot does not account for age-dependent testing patterns.

When the dynamics are projected forward using the inferred parameter and model framework (see Supplementary Methods), we predict an abrupt rise in infections, hospital admissions and deaths (Figure 8) following the rise in S-gene target failure caused by the Omicron variant (Figure 4). In these simulations we include a rapid roll-out of a booster programme vaccinating 6 million people each week, with an expected uptake of 95% in the over 70s, 90% in those aged 50-69 and 80% in those aged 18-50 who have had two doses of vaccine over three months ago. As such we expect the majority of booster vaccination to be complete by early January 2020.

**Figure 8:**
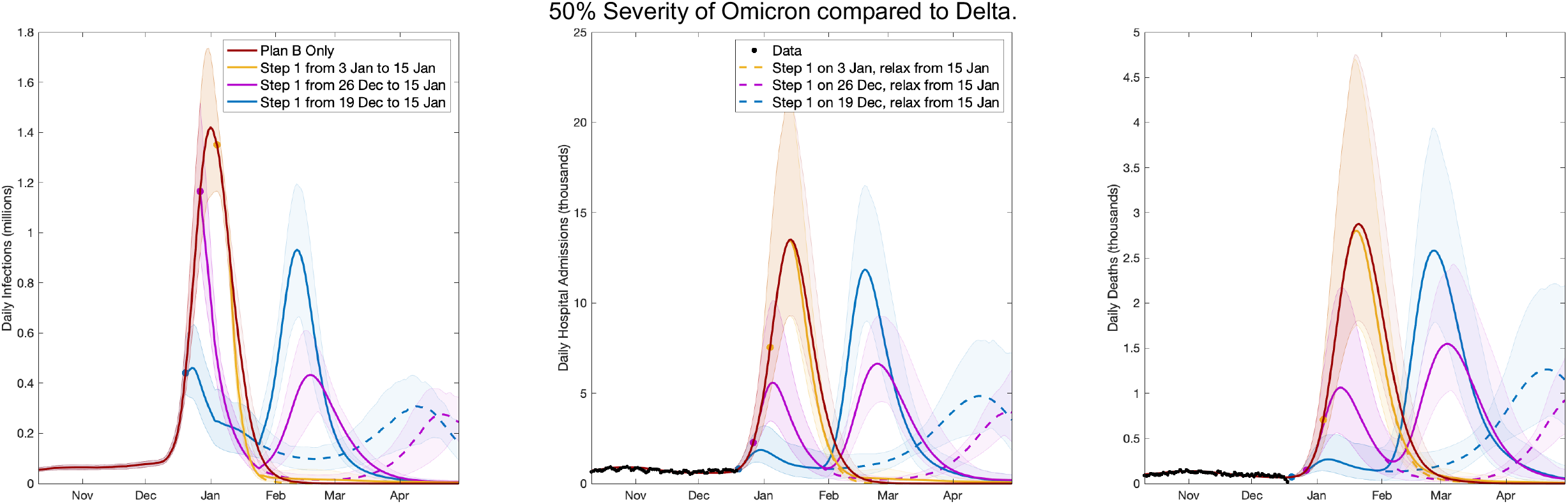
Projected dynamics for infections (left), hospital admissions (centre) and deaths (right), under different control assumptions. The dark red line, which gives the highest peaks, is the mean for the scenario in which only Plan B is enacted; solid lines are where Step 1 type controls are in place from 19th December 2021 (blue), 26th December 2021 (pink) and 3rd January 2022 (yellow) until 15th January 2022 when there is a return to pre-Omicron mixing; dashed lines start Step 1 controls at the same time points, but relaxation starting on 15th January is more gradual. Shaded ribbons correspond to 95% prediction intervals. In this example we assume that Omicron is only 50% as severe as Delta, that is, the associated IHR and IFR (Infection:Hospital ratio and Infection:Fatality ratio) are halved.

We focus initially on simulations were the severity of Omicron is half that of Delta (Figure 8). In the absence of additional controls, with England remaining under Plan B (dark red lines), we project a large wave of infection, leading to hospital admissions peaking at 13,600 per day (9,300-21,3000) and deaths peaking at 2890 per day (1800-4770). The other solid lines (yellow, purple and blue) consider the introduction of a short, stringent circuit-breaker control lasting until 15th January, by which time most people will have received their booster and have the associated protection, after which there is a return to pre-Omicron mixing. In these scenarios, early implementation of controls on 19th or 26th December 2021 (blue or purple) substantially reduces the peaks; however, given the sudden suspension of controls on 15th January 2022, we project an additional wave of infection that could be larger than the initial wave. Later implementation of controls on 3rd January 2022 (yellow) has little impact, with a comparable range of outcomes to the no control scenario.

In practice we are unlikely to see an immediate return to pre-Omicron social behaviour as soon as control measures are lifted. Although Step 4 on 19th July 2021 removed all legal restrictions on social mixing, there was a relative slow drift towards pre-mixing over several months. Similarly, we would not expect the public to instantly transition from a Step-1-type lockdown. We therefore consider scenarios (dashed lines in Figure 8) where Step-1 controls relax over 3 months from 15th January 2022 to 15th April 2022. Under these relaxation assumptions, any resurgence occurs far later and is of lower magnitude, allowing public health services time to consider a range of other mitigation measures.

To investigate the range of potential dynamics more fully, we explore a range of control measures (strength of NPI control and implementation times) and a range of disease severity (from 100% as severe as Delta to just 10%). We primarily focus on hospital admissions under a gradual relaxation of control measures from 15th January (Figure 9), but also consider a rapid return to pre-Omicron measures (Figure S3), and give total infections (Figure S4), hospital bed occupancy (Figure S5) and daily deaths (Figure S6) for a gradual relaxation of control measures.

**Figure 9.**
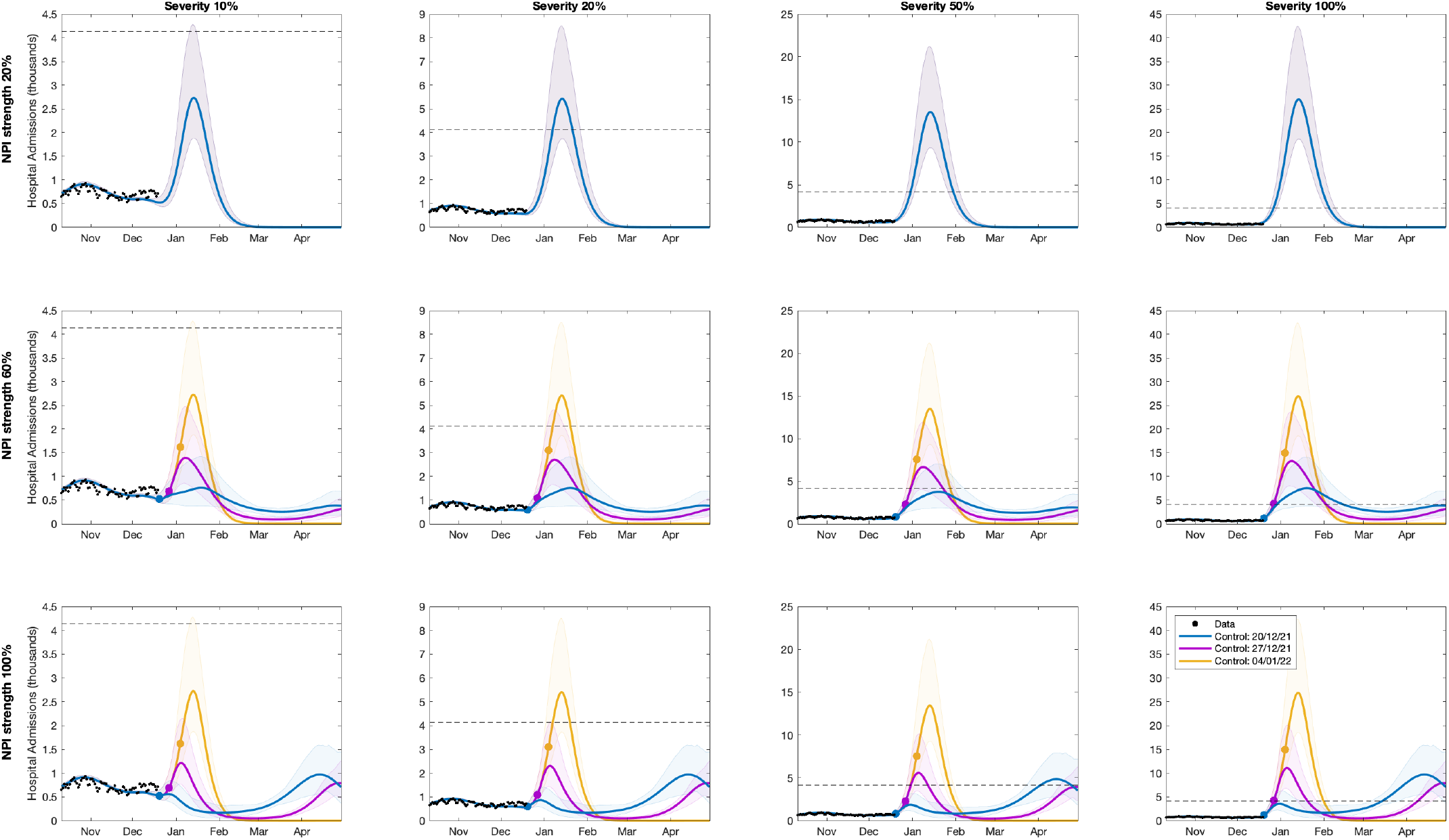
Hospital Admissions for the circuit-breaker model with a gradual return to pre-Omicron mixing from 15th January to 15th April 2022. NPIs of a designated strength are in place from 20th December 2021 (blue), 27th December 2021 (pink) and 4th January 2022 (yellow), indicated by the coloured dots, until 15th January 2022. Solid lines correspond to means and the shaded ribbons the prediction intervals. Black dots represent observed data through to 17th December 2021. The dashed horizontal line corresponds to the previous peak of 4134 admission on 12^th^ January 2021. NPI strengths simulated were (from top row to bottom row): 20% (Plan B with no additional controls), 60%, 100% (Step 1). We also vary the severity of infection with Omicron relative to Delta, scaling the associated IHR and IFR by 10%, 20%, 50% or 100%, respectively (columns, from left to right). Simulations use parameters inferred using data from 17th December.

The top row of each figure refers to an additional 20% control by non-pharmaceutical interventions, compatible with Plan B being maintained from 9th December to 15th January. Under these assumptions of no additional control, and even assuming Omicron is just 10% the severity of Delta (top left of Figure 9) it is still highly likely that hospital admissions will peak above 1500 per day. If we assume that Omicron is as severe as Delta (top right of Figure 9) then admissions will be an order of magnitude larger, peaking at around 27000 admissions (PI 18000-42000).

The middle row increases the strength of short-term control to 60% (approximately comparable with Steps 2 and 3 of the relaxation roadmap - although with the caveat that increasing measures to Step 3 is unlikely to offer the same level of control as when measures reduced to Step 3). This further suppresses the growth of the epidemic but is insufficiently strong to reverse the early growth of Omicron (blue line, Figure S4). Early implementation of such controls (on 19th December) can dramatically suppress the peak number of hospital admissions, even when Omicron is as severe as Delta (middle right of Figure 9), although it produces a long tail of relatively high admissions. However, when Omicron is as severe as Delta (or even half as severe), these control measures still generate a substantial peak as large (if not larger) than previous peaks, placing a huge burden on the health services.

Finally, we consider controls comparable with Step 1 (strong NPI measures but with schools open) in the lower row. Here we see that controls can prevent the rise of Omicron infection (Figure S4), although the hospital admissions continue to rise after controls are implemented due to the delay between infection and the need for hospital treatment. Early implementation (19th December) of these strong control measures leads to a continual decline in hospital admissions over the next two months when Omicron severity is low, and is likely to keep the January 2022 peak just below the scale of the January 2021 peak (4134 on 12th January) for all severities. The suppression of the January 2022 wave means that there is the potential for a large exit wave (although this is lower and more spread out than an uncontrolled wave); however, a delayed exit wave opens the possibility of there being new vaccines available by that time that specifically target the Omicron variant.

## Discussion

Here we have expanded our existing SARS-CoV-2 predictive model to include the emergence and transmission of the Omicron variant. We find evidence of rapid relative growth of the Omicron variant compared to the pre-existing Delta variant in multiple surveillance streams. It is worth stressing that these growth rates are faster than observed for the wild type variant in March 2020, despite the current levels of immunity (from infection and vaccination) and the frequent use of testing. Using these data, we project plausible future scenarios and explore the impact of multiple interventions. While we have a good understanding of the relative growth of Omicron compared to Delta from TaqPath PCR testing, all other measures are far less certain and are strongly influenced by lags within the reporting system and behavioural change.

In many ways, the 100% severity projections represent a reasonable worst case when Omicron has identical disease characteristics to Delta, and the only limits on social mixing come from imposed restrictions. We therefore highlight below six elements that may impact the projected dynamics:

Severity of disease and the vaccine efficacy against severe disease are the pivotal unknown quantities. In all simulations we have explored variations in severity, as characterised by the IHR and IFR (Infection:Hospital ratio, and Infection:Fatality ratio) of Omicron compared to Delta. At the lowest level of severity considered (10%), we only expect a tenth of the severe cases that are projected at the highest level of severity (100%, when Omicron and Delta have the same IFR and IHR values). Changes in severity also help to capture the impact of greater protection for vaccines; our default assumption is that vaccine efficacy against severe disease is 85% following a booster dose, if this value increases to 92.5% then we will halve the number of these individuals requiring hospital admissions - giving a comparable wave to reducing severity to 50%. To estimate hospital occupancy, we assume similar (age-dependent) lengths of stay in hospital for Omicron as for Delta. Shorter lengths of hospital stay for Omicron would reduce hospital occupancies, and thus also reduce pressure on the NHS.

We expect there to have already been considerable behavioural change since Omicron was first reported in the UK the majority of which is yet to be seen in the epidemiological data. Throughout the pandemic we have observed that behaviour does not purely reflect restrictions, but is reactive to perceived risk driven by media reports, general advice and knowledge of local cases. It is highly likely that many of the elderly and most vulnerable have already been taking some measures to limit their exposure. This differential behaviour within the population, would have a limited impact on the general rising levels of infection, but would reduce the numbers that may need to attend hospital. The Warwick model is not partitioned by vulnerability, so this factor is difficult to account for within the projections.

Even if no additional control measures are brought into effect, it is highly likely that the population will continue to change their behaviour in response to increasing levels of infection and presumably increasing media coverage. Such dynamic changes are beyond the current capacity of the model, but would act to reduce the peak -- although the delay from changing behaviour to affecting the admission to hospitals could mean that behaviour changes are too late to have a large effect. Given the high level of testing that is currently undertaken, and the high level of testing expected over the Christmas period, there is also the possibility that some level of control may be afforded by isolation of contacts. Historically, high cases in early July 2021 led to large numbers of individuals isolating - the so-called “pingdemic” - and a dramatic decline in subsequent infections.

Throughout we have assumed that Omicron has the same generation time distribution as Delta - essentially the same latent and infection periods. However, the rapid increase of Omicron relative to Delta could partially be due to a shorter generation time; Omicron would still need to have a competitive advantage over Delta but this would be magnified by a shorter generation time. As such, if the generation time of Omicron was half that of Delta (so around 2.5-3 days instead of approximately 5-6 days), once the model is recalibrated to match the growth of SGTF, this would approximately halve the predicted peak outbreak sizes.

One unknown parameter throughout much of the pandemic has been the impact of vaccination on onward transmission -- the reduction in transmission from a vaccinated individual (compared to a non-vaccinated individual) who becomes infected. We make the assumption that this reduction is 30%, in line with our estimates for Delta, but higher or lower values (either due to the booster or due to lower efficacy against Omicron) will impact the size of the Omicron wave.

The model is deterministic and operates at the scale of NHS regions; both of these factors lead to high levels of spatial and structural synchrony of epidemic waves. Preliminary data suggests that Omicron is concentrated in younger age-groups (20-40) and in urban areas. Less synchrony between age-groups and between spatial locations could lead to multiple asynchronous epidemics in each region, reducing the height of the projected peaks but increasing their duration.

Slight discrepancies in the estimated distribution of lags between infection and detection, or between infection and hospital admission can push the projections out by a few days. Under normal circumstances an error of a few days is irrelevant, but with infection doubling every two days this can substantially disrupt the fit between models and data. It will not substantially change the height of the peak, but can influence the timing.

Despite these caveats, our projections show that Omicron, due to its rapid growth, can generate levels of infection that could disrupt many services and levels of hospital admissions that will place a severe burden on the health services. Determining the optimal control policy is highly dependent on the objective, but several general conclusions can be drawn. First, strong controls enacted early bring the greatest reduction in infections, hospital admissions and deaths during the first wave of Omicron. Second, small initial waves lead to larger exit waves, with exit waves of deaths and hospital admission relatively larger than the exit wave of infection due to changes in the age-distribution of infection. However, such later exit waves, which tend to peak in April 2022, provide the opportunity to learn more about the Omicron variant and to instigate specific controls.

## Data Availability

Data on reported cases, deaths and vaccination used in this model are at the individual level and therefore confidential, with public data deposition non-permissible for socioeconomic reasons. The ethics of the use of these data for these purposes was agreed by the UK Health Security Agency with the Governments SPI-M(O) / SAGE committees. More aggregate data is freely available from the UK Coronavirus dashboard: https://coronavirus.data.gov.uk/

## SUPPLEMENTARY FIGURES

**Figure S1.**
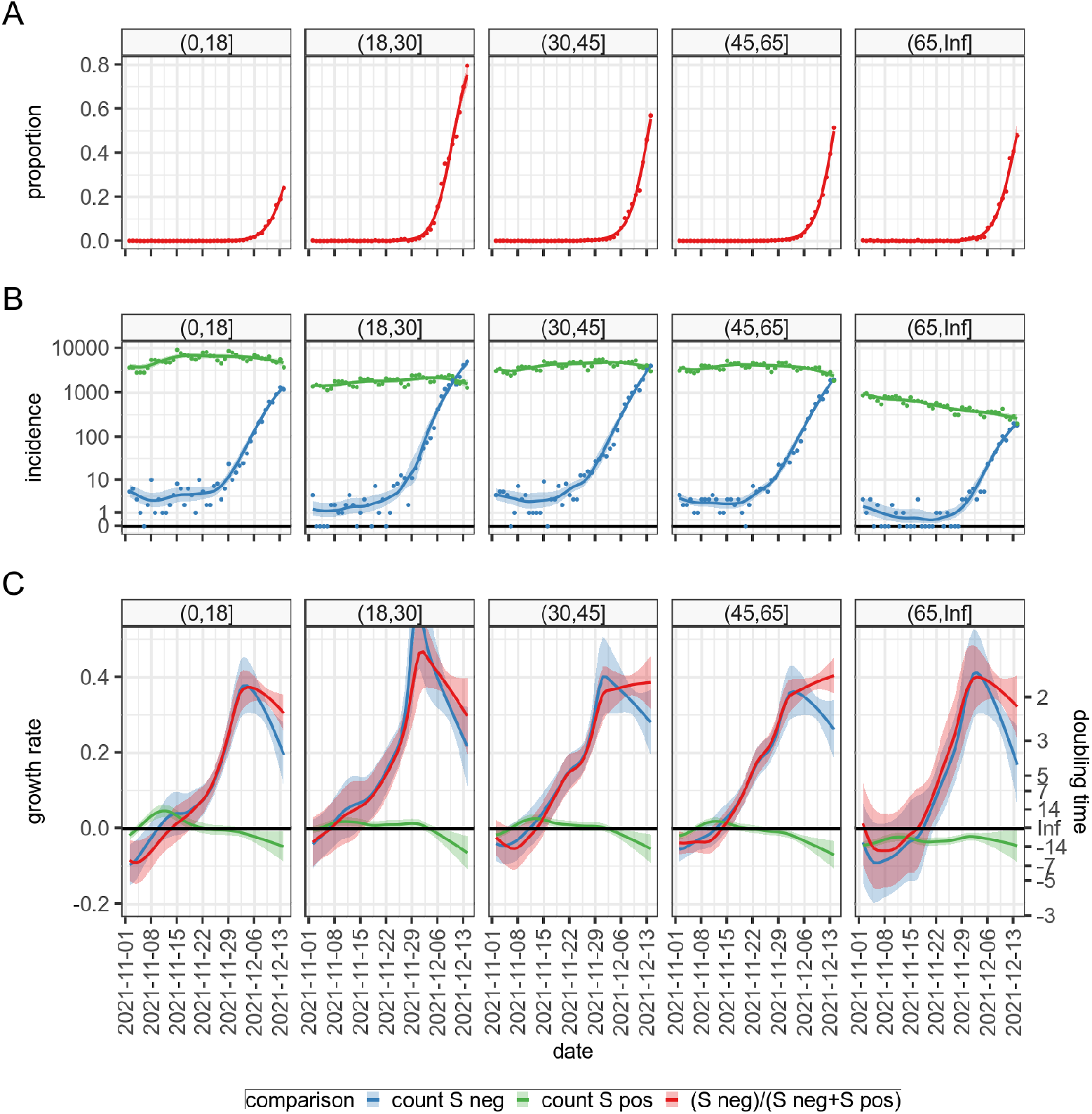
Dynamics of S-gene target failure (blue), S-gene detection (green) and the ratio (red) for five age-ranges in England. Although those 18-30 show the greatest proportion of S-gene target failures, and hence the greatest assumed proportion of Omicron variant, all age-groups show comparable patterns of growth. This suggests that although Omicron was initially concentrated in the 18-30 age group, it has now spread to all others and is growing at a similar rate. Similar findings are observed for different UK regions, and ethnicities (not shown).

**Figure S2.**
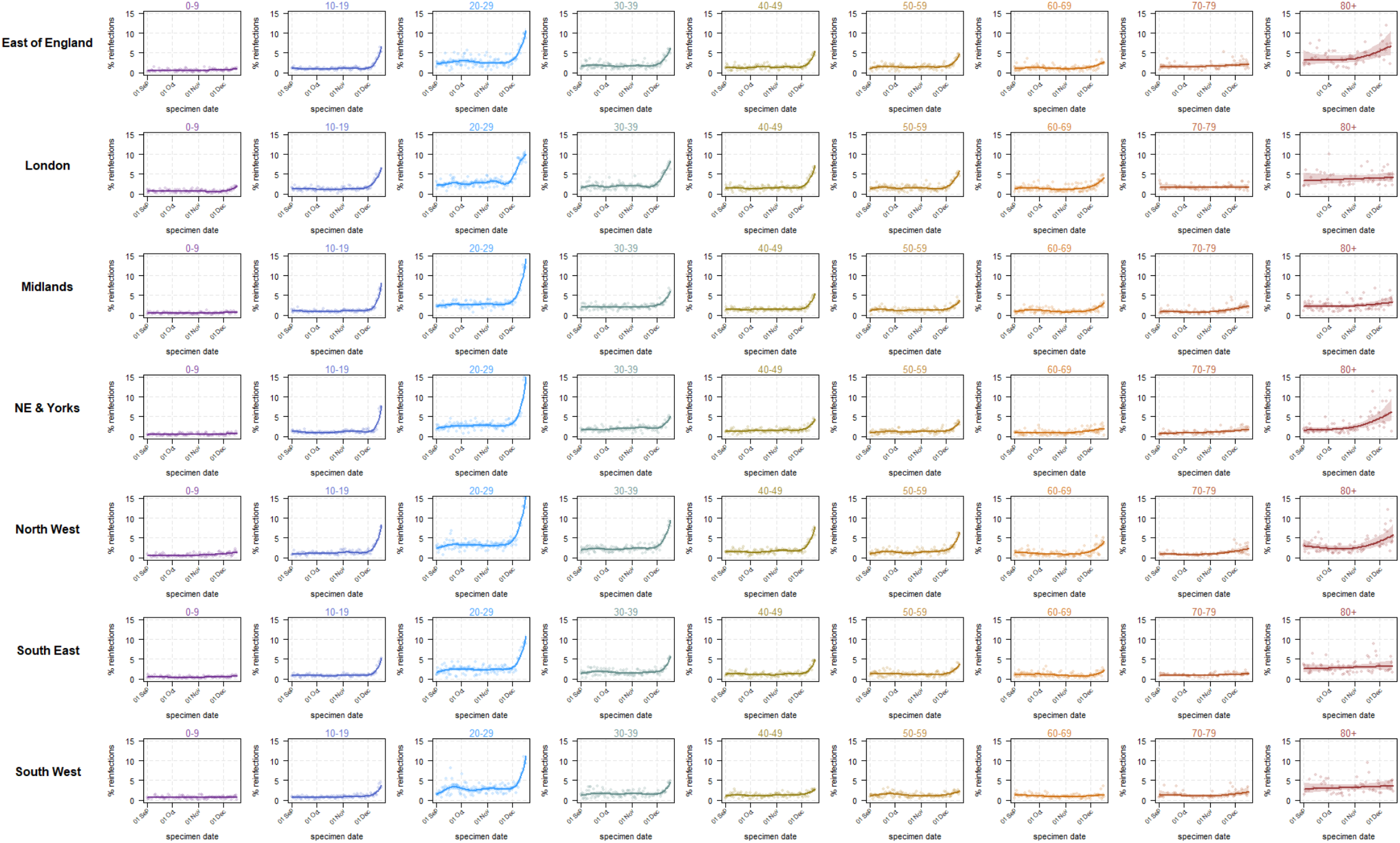
Estimated daily % of cases that are reinfections for England, stratified by age and region.Observations are shown as dots, fitted splines as lines and the 95% confidence intervals as shaded regions.

**Figure S3.**
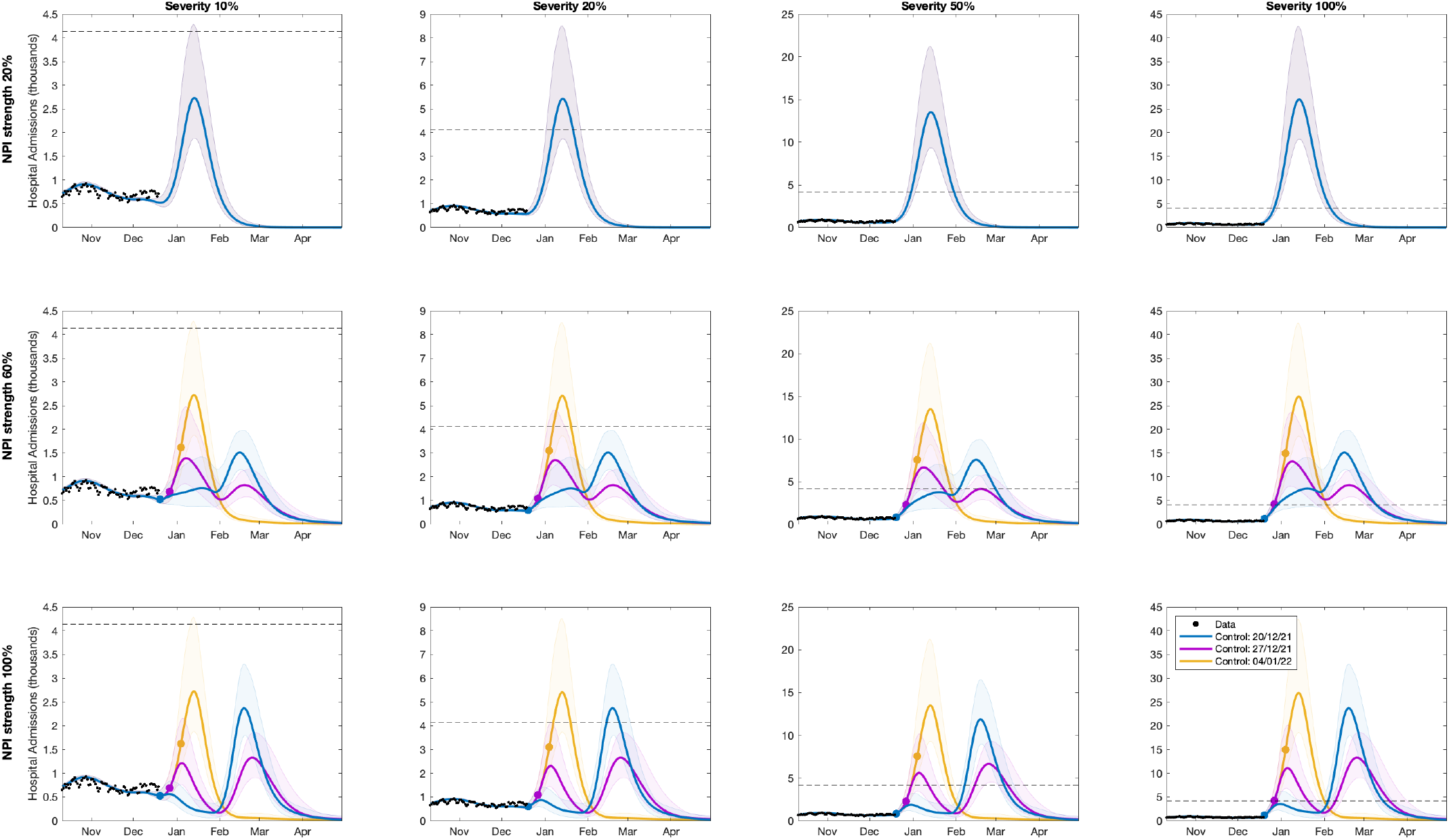
Hospital Admissions for the circuit-breaker model with a rapid return to pre-Omicron mixing on 15th January 2022. NPIs of a designated strength are in place from 20th December 2021 (blue), 27th December 2021 (pink) and 4th January 2022 (yellow), indicated by the coloured dots, until 15th January 2022. Solid lines correspond to means and the shaded ribbons the prediction intervals. Black dots represent observed data through to 17th December 2021. The horizontal line represents the previous peak of 4134 admissions on 12^th^ January 2021. NPI strengths simulated were (from top row to bottom row): 20% (Plan B with no additional controls), 60%, 100% (Step 1). We also vary the severity of infection with Omicron relative to Delta, scaling the associated IHR and IFR by 10%, 20%, 50% or 100%, respectively (columns, from left to right). Simulations use parameters inferred using data from 17th December.

**Figure S4.**
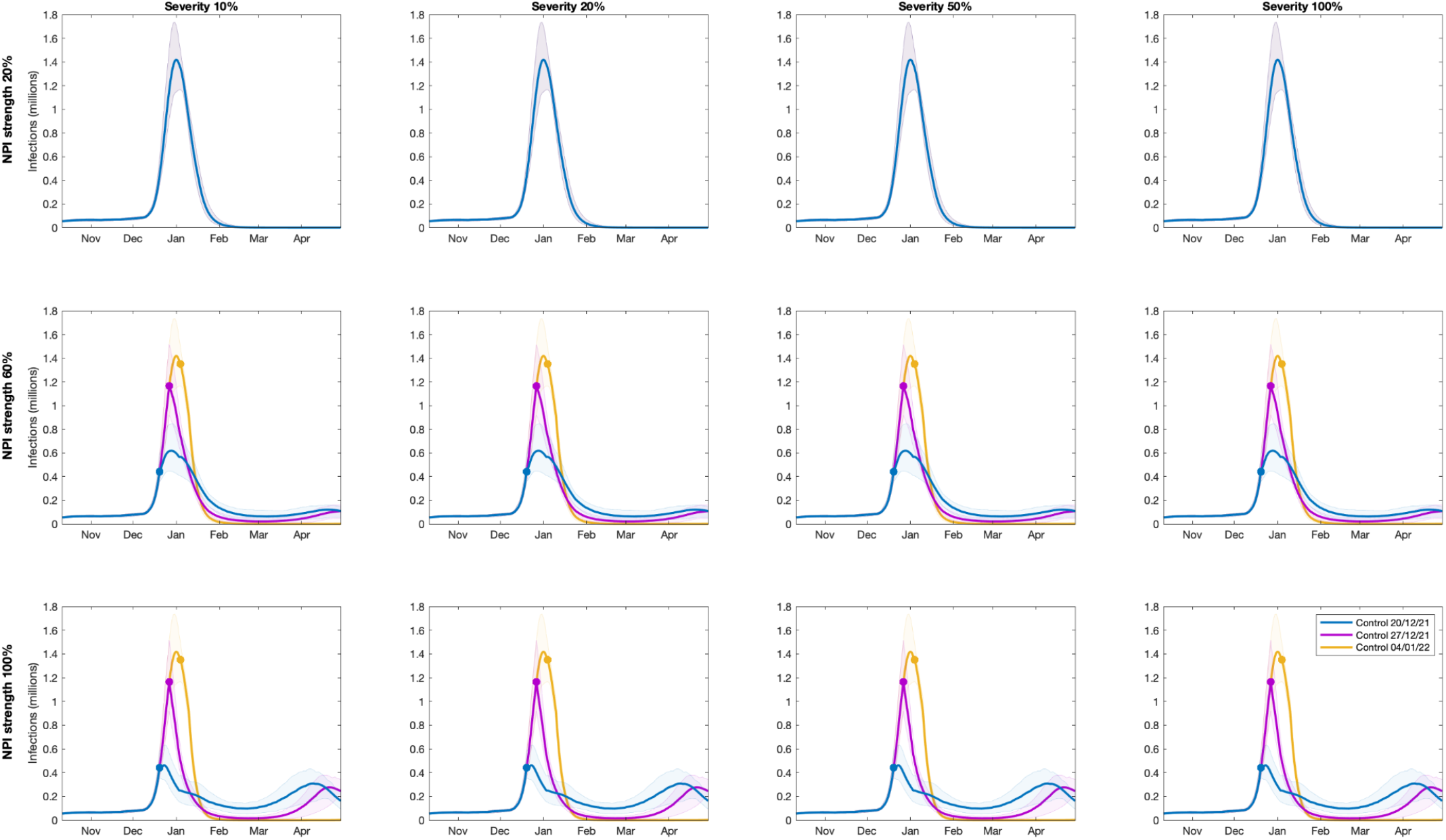
All new infections (symptomatic, asymptomatic, first infections and re-infections) for the circuit-breaker model with a gradual return to pre-Omicron mixing from 15th January to 15th April 2022. NPIs of a designated strength are in place from 20th December 2021 (blue), 27th December 2021 (pink) and 4th January 2022 (yellow), indicated by the coloured dots, until 15th January 2022. Solid lines correspond to means and the shaded ribbons the prediction intervals. NPI strengths simulated were (from top row to bottom row): 20% (Plan B with no additional controls), 60%, 100% (Step 1). We also vary the severity of infection with Omicron relative to Delta, scaling the associated IHR and IFR by 10%, 20%, 50% or 100%, respectively (columns, from left to right); it is assumed that length of stay is the same for Omicron and Delta. Simulations use parameters inferred using data from 17th December.

**Figure S5.**
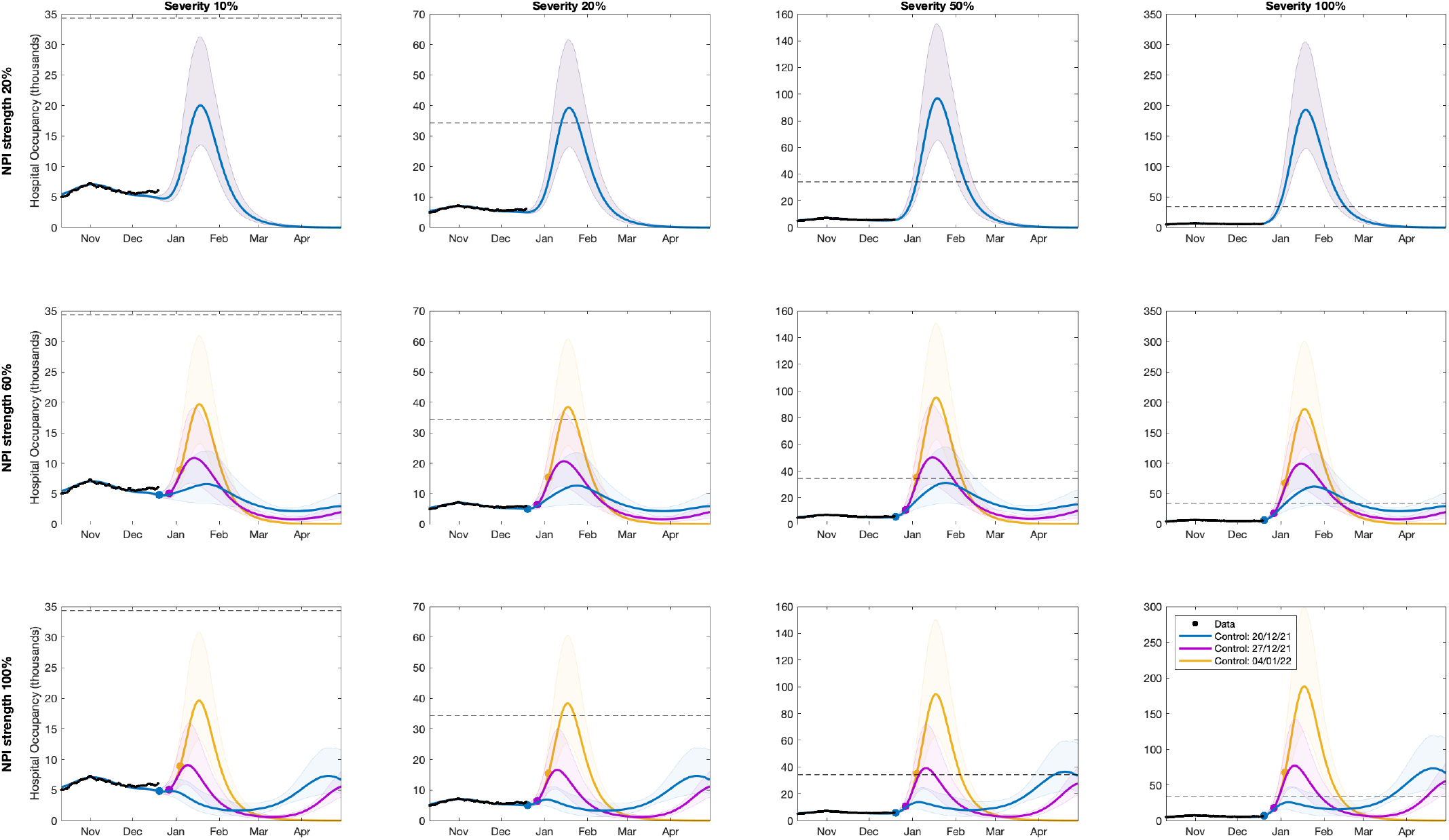
Hospital Bed Occupancy for the circuit-breaker model with a gradual return to pre-Omicron mixing from 15th January to 15th April 2022. NPIs of a designated strength are in place from 20th December 2021 (blue), 27th December 2021 (pink) and 4th January 2022 (yellow) until 15th January 2022. Solid lines correspond to means and the shaded ribbons the prediction intervals. Black dots represent observed data through to 17th December 2021. The horizontal line represents the previous peak occupancy of 34,336 on 18^th^ January 2021. NPI strengths simulated were (from top row to bottom row): 20% (Plan B with no additional controls), 60%, 100% (Step 1). We also vary the severity of infection with Omicron relative to Delta, scaling the associated IHR and IFR by 10%, 20%, 50% or 100%, respectively (columns, from left to right); it is assumed that length of stay is the same for Omicron and Delta. Simulations use parameters inferred using data from 17th December.

**Figure S6.**
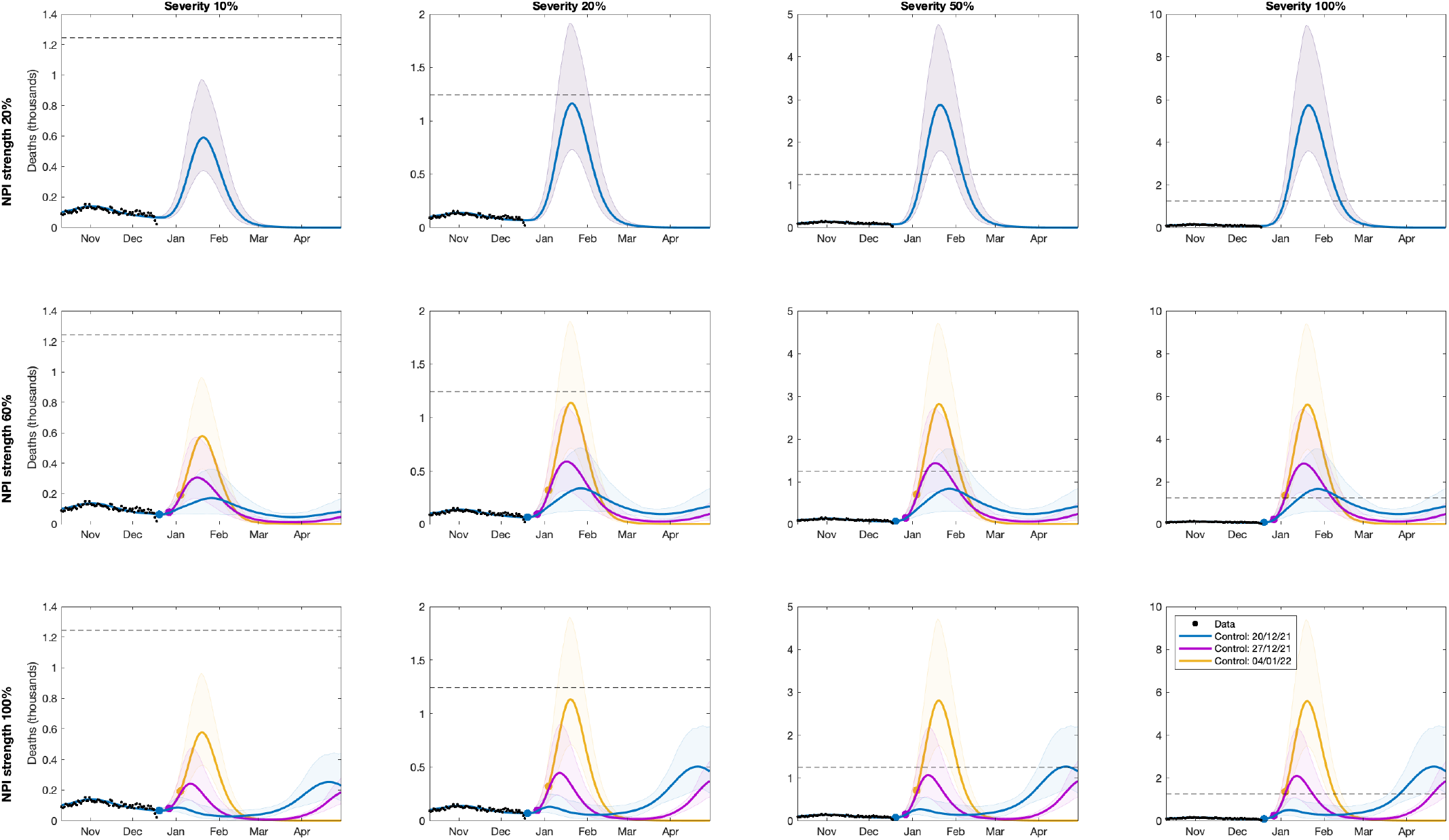
Daily deaths for the circuit-breaker model with a gradual return to pre-Omicron mixing from 15th January to 15th April 2022. NPIs of a designated strength are in place from 20th December 2021 (blue), 27th December 2021 (pink) and 4th January 2022 (yellow) until 15th January 2022. Solid lines correspond to means and the shaded ribbons the prediction intervals. Black dots represent observed data through to 17th December 2021. The horizontal line represents the previous peak of 1244 on 19^th^ January 2021 NPI strengths simulated were (from top row to bottom row): 20% (Plan B with no additional controls), 60%, 100% (Step 1). We also vary the severity of infection with Omicron relative to Delta, scaling the associated IHR and IFR by 10%, 20%, 50% or 100%, respectively (columns, from left to right). Simulations use parameters inferred using data from 17th December.

## SUPPLEMENTARY METHODS

Here we detail the underlying mathematical framework that defines the transmission model. We break the model into multiple sections that combine to generate a picture of SARS-CoV-2 transmission in the UK. This model structure has been detailed in previous publications [14,15,19-21] but we review the details here for completeness.

### Infection modelling

As is common to most epidemiological modelling we stratify the population into multiple disjoint compartments and capture the flow of the population between compartments in terms of ordinary differential equations. At the heart of the model is a modified SEIR equation, where individuals may be susceptible (S), exposed (E), infectious with symptoms (I), infectious and either asymptomatic or with very mild symptoms (A) or recovered (R). Both symptomatic and asymptomatic individuals are able to transmit infection, but asymptomatic infections do so at a reduced rate given by *τ*. Hence, the force of infection is proportional to I+*τ*A. To some extent, the separation into symptomatic (I) and asymptomatic (A) states within the model is somewhat artificial as there are a wide spectrum of symptom severities that can be experienced, with the classification of symptoms changing over time. Our classification reflects early case detection, when only relatively severe symptoms were recognised.

To obtain a better match to the infection time scales, we model the exposed class as a 3-stage process - this provides a better match to the time from infection to becoming infectious, such that in a stochastic formulation the distribution of the latent period would be an Erlang distribution.

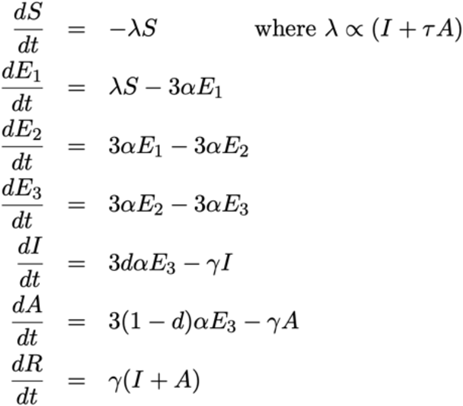

where *α*^-1^, and *γ*^-1^ are the mean latent and infectious periods, while *d* is the proportion of infections that develop symptoms.

### Age Structure & Transmission Structure

The simple model structure is expanded to twenty-one 5-year age-groups (0-4, 5-9,, 95-99, 100+). Age has three major impacts on the epidemiological dynamics, with each element parameterised from the available data:

- Older individuals have a higher susceptibility to SARS-CoV-2 infection (captured by the parameter *σ*).
- Older individuals have a higher risk of developing symptoms, and therefore have a greater rate of transmission per contact.
- Older individuals have a higher risk of more severe consequences of infection including hospital admission and death.

The age-groups interact through four who-acquired-infection-from-whom transmission matrices, which capture the epidemiological relevant mixing in four settings: household (***β***^H^), school (***β***^S^), workplace (***β***^W^) and other (***β***^O^). We took these matrices from Prem *et al*. [22] to allow easy translation to other geographic settings, although other sources could be used.

One of the main modifiers of mixing and therefore transmission is the level of precautionary behaviour, *ϕ* (see Figure 2 of the main text). This scaling parameter changes the who-acquired-infection-from-whom transmission matrices in each transmission setting, such that when *ϕ*=1 mixing in workplaces and other settings take their lowest value, whereas when *ϕ*=0 the mixing returns to pre-pandemic levels. Mixing within the school setting follows the prescribed opening and closing of schools.

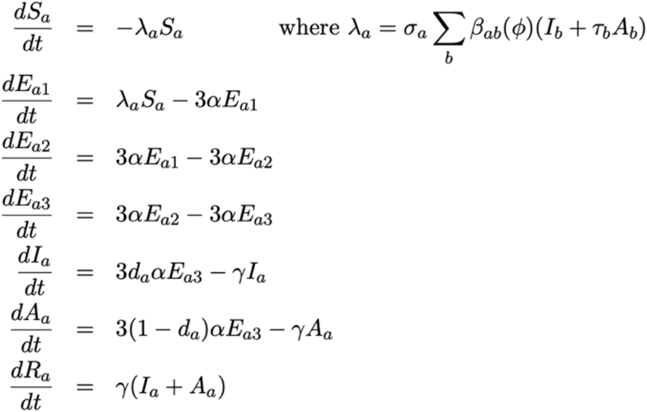

For simplicity of notation, we write the sum of the four age-structured mixing matrices as ***β***(*ϕ*).

To ensure that we can replicate the long-term dynamics of infection we allow the population to age. The aging process occurs annually (corresponding to the new school year in September) in which approximately one fifth of each age-group moves to the next oldest age cohort — small changes to the proportion moving between age-groups are made to keep the population size within each age-group constant.

### Capturing Quarantining

One of the key characteristics of the COVID-19 pandemic in the UK has been the use of self-isolation and household quarantining to reduce transmission. We approximate this process by distinguishing between first infections (caused by infection related to any non-household mixing) and subsequent household infections (caused by infection due to household mixing). The first symptomatic case within a household (which might not be the first infection) has a probability (*H*) of leading to household quarantining; this curtails the non-household mixing of the individual and all subsequent infections generated by this individual.

In our notation, we let superscripts denote the first infection in a household (F), a subsequent infection from a symptomatic household member (SI) and a subsequent infection from an asymptomatic household member (SA); the first detected case in a household who is quarantined (QF) and all their subsequent household infections (QS). For a simple SEIR model (ignoring multiple E categories and age-structure) our extension would give:

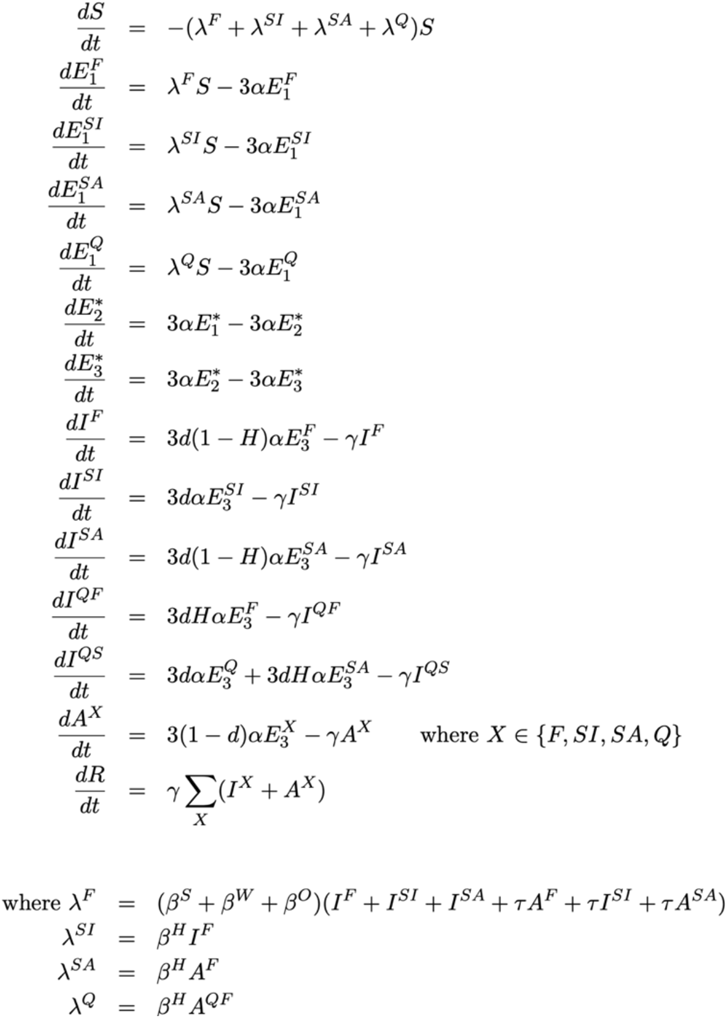

This formation has been shown to be able to reduce *R* below one even when there is strong within household transmission, as infection from quarantined individuals cannot escape the household [19].

### Spatial Modelling

Within England the model operates at the scale of NHS regions (East of England, London, Midlands, North East, North West, South East and South West). For simplicity and speed of simulation we assume that each of these regions acts independently and in isolation - we do not model the movement of people or infection across borders. In addition, the majority of parameters are regionally specific, reflecting different demographics, deprivation and social structures within each region. However, we include a hyper-prior on the shared parameters such that the behaviour of each region helps inform the value in others.

### Variant Modelling

The model also captures the three main variants that have been responsible for most infections in England: the wildtype virus (encapsulating all pre-Alpha variants), the Alpha variant and the Delta variant. Each of these requires a replication of the infectious states for each variant type modelled. We assume that infection with each variant confers immunity to all variants, such that there is indirect competition for susceptible individuals. This competition is driven by the transmission advantage of each variant which is estimated by matching to the proportion of positive community PCR tests (Pillar 2 test) that are positive for the S-gene. The TaqPath system that is used for the majority of PCR tests in England is unable to detect the S-gene in Alpha variants, due to mutations in the S-gene. The switch from S-gene positive to S-gene negative and back to S-gene positive corresponds with the dominance of wildtype, Alpha and Delta variants. We infer the transmissibility of Alpha and Delta variants to be 52% (35-71%) and 156% (117-210%) greater than wildtype, respectively. It is into this structure that we include the Omicron variant, setting the vaccine efficacy and level of cross protection, and varying the transmission rate to capture the rapid increase in infection.

### Vaccination Modelling

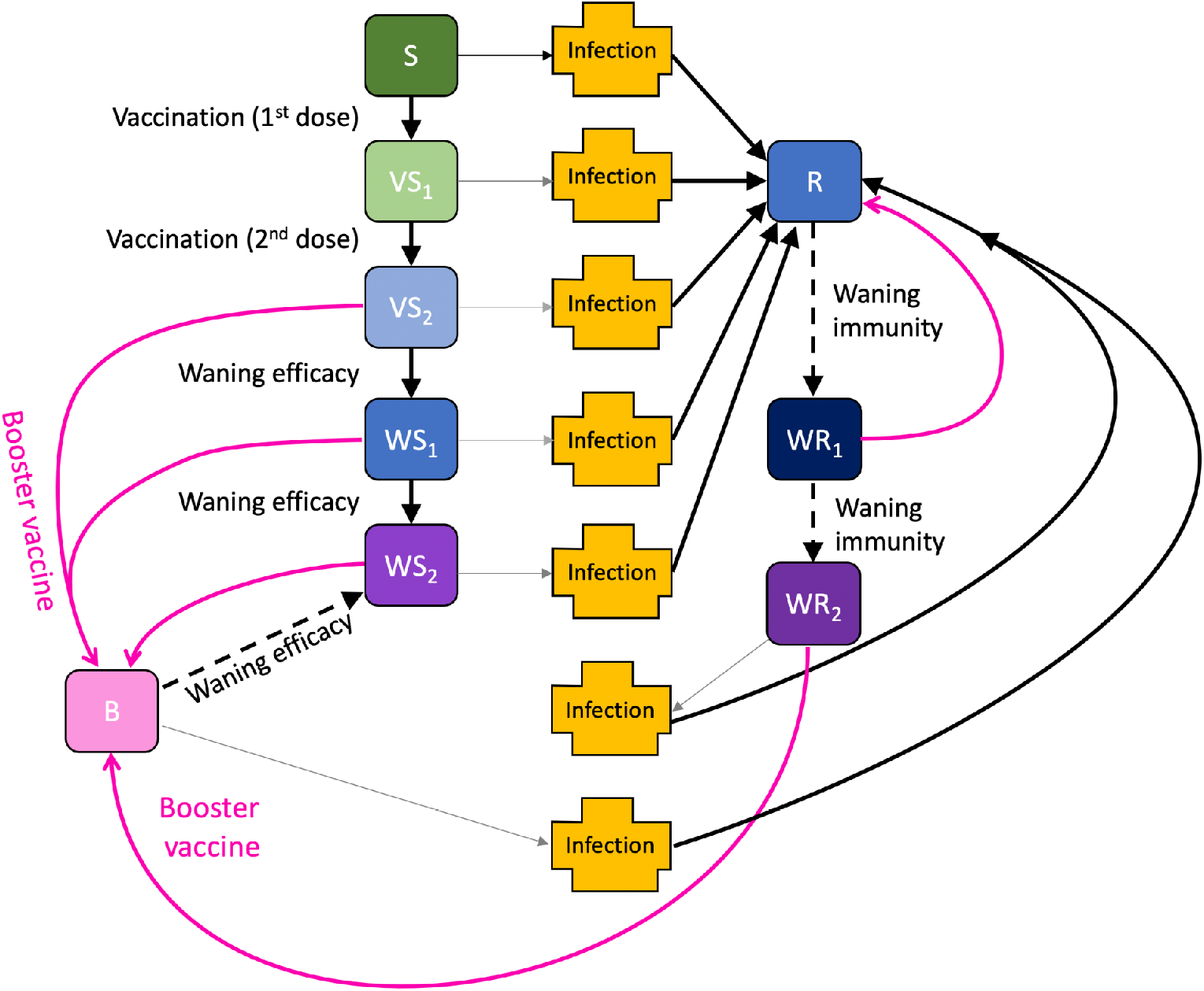

We capture vaccination using a leaky approach, although non-leaky (all-or-nothing) models produce extremely similar results over the time-scales considered. The model replicates the action of:

- first and second doses of vaccine, at rates *v*_1_ and *v*_2_ respectively that move susceptible individuals through to vaccinated states (V*S*_1_ and V*S*_2_) but have no impact on infected or recovered individuals;
- waning vaccine efficacy at rates *ω*_1_ and *ω*_2_, giving a two-step process from fully vaccinated to waned efficacy (in the equation below, for simplicity we assume everyone who gets a first dose of vaccine also gets a second, so that waning from state *S*_1_ is unnecessary);
- waning immunity at rates *Ω*_1_ and *Ω*_2_ which are assumed to be slower than the waning of vaccine efficacy.

The model also needs to capture the total number of individuals who have been given a first or second dose of vaccine (*V*_1_ or *V*_2_ out of a total population size of *N*) to ensure that only individuals that have not been vaccinated are offered a first dose, and only individuals that have been vaccinated once are offered a second dose.

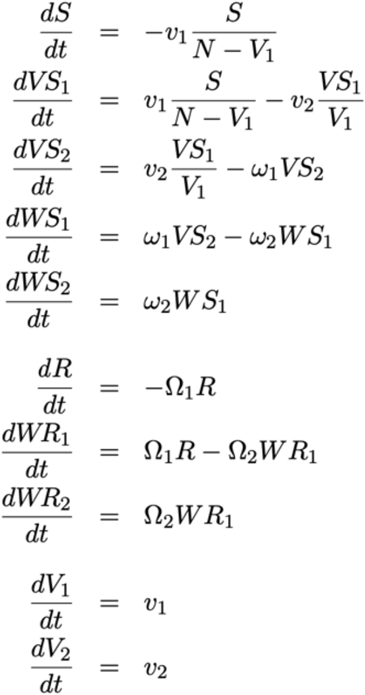

For those in the classes where the vaccines generate protection (*VS*_1_, *VS*_2_ and *WS*_1_), the degree of protection is determined by the ratio of AstraZeneca (ChAdOx1) vaccine to mRNA vaccines (either Pfizer BNT162b2 or the Moderna COVID-19 vaccine) that has been given to that age group (see Table 1). If a vaccinated individual becomes infected, their probability of being admitted to hospital or dying - which normally only depends on age - is modified by the appropriate vaccine efficacy according to the ratio of the two vaccine types. Booster vaccinations are implemented by moving individuals in the vaccinated or waned class into a booster class where the level of protection is enhanced. Waning from the booster class occurs at a low rate.

**Table 1.** Vaccine efficacy and protection from prior infection assumptions for the Delta variant and Omicron variant. In England, individuals who receive COVID-19 booster vaccinations receive a full dose of Pfizer or a half dose of Moderna, regardless of which primary course was received. AZ = ChAdOx1 vaccine; mRNA = BNT162b2 (Pfizer) or mRNA-1273 (Moderna) vaccines; 1 = one dose; 2 = two doses; B = booster dose, W = waned protection, and mean time to reach that value.

| Variant | Outcome | Prior infection |  |  | AZ primary, mRNA booster |  |  |  | mRNA primary, mRNA booster |  |  |  |
| --- | --- | --- | --- | --- | --- | --- | --- | --- | --- | --- | --- | --- |
|  |  | Non-Omicron | Omicron | W | 1 | 2 | B | W | 1 | 2 | B | W |
| Delta | Infection | 100% | 100% | 0%,<br>1860 days | 45% | 70% | 88% | 0%,<br>460 days | 55% | 85% | 88% | 0%,<br>460 days |
|  | Symptoms | 100% | 100% | 0%,<br>1860 days | 45% | 70% | 92% | 0%,<br>460 days | 55% | 90% | 92% | 0%,<br>460 days |
|  | Hospital Adm. | 100% | 100% | 0%,<br>1860 days | 80% | 92% | 96% | 0%,<br>460 days | 80% | 95% | 96% | 0%,<br>460 days |
|  | Death | 100% | 100% | 0%,<br>1860 days | 80% | 97% | 99% | 0%,<br>460 days | 80% | 98% | 99% | 0%,<br>460 days |
| Omicron | Infection | 90% | 100% | 0%,<br>1860 days | 4% | 24% | 60% | 0%,<br>460 days | 9% | 52% | 60% | 0%,<br>460 days |
|  | Symptoms | 90% | 100% | 0%,<br>1860 days | 4% | 24% | 72% | 0%,<br>460 days | 9% | 52% | 72% | 0%,<br>460 days |
|  | Hospital Adm. | 90% | 100% | 0%,<br>1860 days | 41% | 71% | 85% | 0%,<br>460 days | 41% | 81% | 85% | 0%,<br>460 days |
|  | Death | 90% | 100% | 0%,<br>1860 days | 41% | 88% | 94% | 0%,<br>460 days | 41% | 92% | 94% | 0%,<br>460 days |

### Parameter Inference

Key to the accuracy of any model are the parameters that underpin the dynamics. With a model of this complexity, a large number of parameters are required. Some, such as vaccine efficacy, are assumed values based on the current literature; while others, such as the level of precautionary behaviour over time, are inferred from the epidemic dynamics. Bayesian inference, using an MCMC process, is applied to each of the seven NHS regions in England to determine posterior distributions for each of the regional parameters (further details are given in [14]). The distribution of parameters leads to uncertainty in model projections, which is represented by the 95% prediction interval in all graphs (this interval contains 95% of all predictions).

As the epidemic has progressed, new posterior distributions based on the latest data are initialised from previous MCMC chains – ensuring a rapid fit to historical data. We match to six observations: hospital admissions, hospital occupancy, ICU occupancy, deaths, proportion of pillar 2 (community) test that are position, and the proportion of pillar 2 tests that are S-gene positive (as a signal of the ratio of wild-type to Alpha variant, then a signal of the ratio of Delta to Alpha variant, and more recently a signal of Omicron to Delta). Although not part of the transmission dynamics, these six quantities for each region can be generated from the number, age and type of infection within the model. Observations and model results are compared by considering the likelihood of generating the observations assuming they are Poisson distributed with a mean given by the results of the deterministic model.

## Data Availability

Data on cases were obtained from the COVID-19 Hospitalisation in England Surveillance System (CHESS) data set that collects detailed data on patients infected with COVID-19. Data on COVID-19 deaths were obtained from Public Health England. These data contain confidential information, with public data deposition non-permissible for socioeconomic reasons. The CHESS data resides with the National Health Service (www.nhs.gov.uk) whilst the death data are available from Public Health England (www.phe.gov.uk).

## Patient and public involvement

This was an urgent public health research study in response to a Public Health Emergency of International Concern. Patients or the public were not involved in the design, conduct, or reporting of this rapid response research.

## Ethical considerations

Data from the CHESS database were supplied after anonymisation under strict data protection protocols agreed between the University of Warwick and Public Health England. The ethics of the use of these data for these purposes was agreed by Public Health England with the Government’s SPI-M(O) / SAGE committees.

